# Identifying STEMI Patients for Early Intensive Care Unit Discharge: The Codi-IAM Score

**DOI:** 10.64898/2025.12.08.25341864

**Authors:** Giovanni Occhipinti, Daniel Vilar-Roquet, Jordi Guarinos Oltra, Juan Andrés Bermeo Garrido, Xavier Carrillo Suarez, Marcelo Jimenez Kockar, Laia Milà, Rut Andrea Riba, Merida Cardenas Manilla, Juan Francisco Muñoz Camacho, Carlos Tomas-Querol, Nuria Ribas Barquet, Mohsen Mohandes Yusefian, Irene Buera Surribas, Cosme García, Oriol de Diego, Cinta Llibre Pallares, Albert Ariza, Xavier Freixa, José Antonio Barrabés, Ander Regueiro, Josep Rodes-Cabau, Manel Sabaté Tenas, Josepa Mauri Ferre, Helena Tizón-Marcos, Omar Abdul-Jawad Altisent

**Affiliations:** Institut Clinic Cardiovascular, Hospital Clínic de Barcelona, University of Barcelona, Barcelona, Spain; Instituto de Investigaciones Biomédicas Agusti Pi i Sunyer (IDIBAPS), Barcelona, Spain; Area Assistencial CatSalut, Barcelona, Spain; Hospital Universitari Joan XXIII, Tarragona, Spain; Hospital Universitari de Bellvitge, Hospitalet de Llobregat, Spain; Hospital Universitari Germas Trias, Barcelona, Spain; Hospital de la Santa Creu i Sant Pau, Barcelona, Spain; Hospital de la Vall d’Hebron, Barcelona, Spain; Hospital Josep Trueta, Girona, Spain; Hospital MutuaTerrassa, Terrassa, Spain; Hospital Arnau de Vilanova, Lleida, Spain; Hospital del Mar, Barcelona, Spain; Hospital Laval, Quebec, Canada; Centro de Investogación Biomédica en Red. Enfermedades Cardiovasculares (CIBER-CV)

**Keywords:** Catheterization, coronary angiography, coronary artery disease, STEMI, percutaneous coronary Intervention, Early discharge

## Abstract

**Background:** Prolonged intensive care unit (ICU) monitoring after primary percutaneous coronary intervention (pPCI) for ST-segment elevation myocardial infarction (STEMI) may be unnecessary for truly low-risk patients, yet no bedside tool reliably classify these patients. We aimed to develop and externally validate a score that identifies STEMI patients at very low risk of 30-day death or ICU-level complications after pPCI and to examine its prognostic value through two years.

**Methods:** From July 2020 to July 2022, 5,093 consecutive STEMI patients treated with pPCI ≤24 hours from symptom onset were prospectively enrolled across 10 centers in Catalonia Region. Derivation (n = 3,649) and validation (n = 1,444) cohorts were pre-specified. Multivariable logistic regression isolated independent predictors of the composite endpoint (30-day all-cause death or in-hospital complications requiring ICU care). Each predictor was weighted by its odds ratio to generate the Codi-IAM Score (0–24 points).

**Results:** Seven variables entered the score: age > 75 years, Killip > I, malignant peri-procedural arrhythmia, left-main disease, three-vessel disease, non-radial access, and unsuccessful PCI (final TIMI < 3). Discrimination was excellent (AUC 0.87 derivation; 0.86 validation). Very low-risk patients (score 0; 44.3% of the population) experienced no deaths within 72 hours, a 1.7% in-hospital complication rate, and 0.2% 30-day mortality; two-year survival exceeded 98%.

**Conclusions:** The Codi-IAM Score, available immediately after pPCI, accurately flags nearly half of STEMI patients as having negligible early risk. Its adoption could safely shorten ICU monitoring and hospital stay, improving resource allocation without impact on clinical outcomes.

**What Is Known:**

- Early intensive care unit (ICU) discharge after primary percutaneous coronary intervention (pPCI) for ST-segment elevation myocardial infarction (STEMI) is recommended by current guidelines but remains limited by safety concerns.
- Current European Society of Cardiology guidelines recommend at least 24 hours of continuous ICU monitoring for all STEMI patients, with only class IIb recommendation for early discharge within 72 hours.
- Prolonged ICU stays may be associated with increased nosocomial complications, delayed mobilization, higher costs, and inefficient use of critical care resources, particularly relevant during healthcare system strain.

**What the Study Adds:**

- The Codi-IAM Score is a simple, immediately available bedside tool incorporating seven variables that accurately identifies STEMI patients at very low risk of 30-day mortality or ICU-level complications after pPCI.
- Nearly half of STEMI patients (44.3%) score 0 on the Codi-IAM Score and experience negligible early risk: no deaths within 72 hours, 1.7% in-hospital complication rate, and 0.2% 30-day mortality.
- Implementation of the Codi-IAM Score in clinical practice could safely shorten ICU stays and hospital length of stay in approximately half of STEMI patients, improving resource allocation and patient satisfaction without compromising clinical outcomes over two-year follow-up.

## INTRODUCTION

Early intensive care unit (ICU) discharge following primary percutaneous coronary intervention (pPCI) for ST-segment elevation myocardial infarction (STEMI) is gaining broader acceptance. This strategy is well-perceived by patients and offers the potential to streamline care delivery and reduce healthcare resource utilization by shortening hospital stays.(1, 2, 3) However, its widespread adoption remains limited due to persistent safety concerns, primarily stemming from the lack of validated, bedside, risk stratification tools capable of reliably identifying patients at minimal risk for early post-procedural complications.

Current European Society of Cardiology guidelines recommend at least 24 hours of continuous intensive care unit (ICU) monitoring for all STEMI patients, regardless of their risk profile, with only a class IIb recommendation supporting early discharge within 72 hours.(4) American guidelines do not provide a specific recommendation regarding ICU monitoring duration.(5) Consequently, ICU care lasting 24 hours or more remains the standard practice. Yet, prolonged ICU stays may be associated with increased nosocomial risk, delayed patient mobilization, higher costs, and inefficient use of critical care resources, challenges that were particularly pronounced during the COVID-19 pandemic.(6, 7) Emerging data suggest that a well-defined subgroup of STEMI patients, characterized by preserved hemodynamic, successful revascularization, and uncomplicated procedural courses, has a negligible risk of major adverse events beyond the initial 24 hours post-pPCI.(1) When paired with structured multidisciplinary follow-up, early discharge in this population may yield equal or improved outcomes compared to standard care.(8)

To address the current gap in pragmatic, real time assessment, we conducted a multicenter study with two primary objectives: i) to externally validate our previously identified predictors of very low risk among STEMI patients treated with pPCI;(9) and ii) to develop a simple, clinical applicable bedside risk score to guide safe and efficient early ICU discharge decisions.

## METHODS

### Study Design and Population

This was a prospective, multicenter study including consecutive patients with a presumptive diagnosis of STEMI enrolled through the Catalonia Acute Myocardial Infarction (AMI)-Code network between July 1, 2020, and July 31, 2022. The Catalonia AMI-Code is a regional STEMI care system including 10 hospitals offering 24/7 pPCI coverage for ∼8 million inhabitants. The structure, logistics, and operational protocols of the network have been described elsewhere and are further detailed in the **Supplementary Materials** (**Supplementary Methods Section**).(10) Briefly, patients presenting with chest pain are triaged via clinical assessment and 12-lead ECG by emergency medical services (EMS), or at non-PCI-capable hospitals. When STEMI is suspected, the AMI-Code is activated to facilitate rapid transfer to the nearest PCI-capable center through a centralized logistics hub. Throughout the AMI-Code network, a total of 5,918 AMI-code activations were identified. Among those, patients with alternative diagnoses (i.e., false activation; n=277), those treated with fibrinolysis (n=6), and those presenting >24 hours after symptom onset (n=543) were excluded. The final study cohort consisted of 5,093 confirmed STEMI patients undergoing pPCI within 24 hours from symptoms onset (**Supplementary Figure 1**).

### Data Collection and Definitions

Clinical and procedural data were prospectively collected in the “Codi-IAM” online registry, where data are recorded by trained physicians at the time of the index event. The registry includes automated data validation checks, completeness monitoring, routine on-site audits, and annual data quality reviews by the Catalonia Department of Public Health, to guarantee integrity and accuracy of registry data (**Supplementary Materials, Supplementary Methods Section**). Collected variables included demographics, comorbidities, clinical presentation, procedural details, and timing intervals (e.g., first medical contact [FMC] and door-to-balloon times). Successful PCI was defined as achieving final Thrombolysis in Myocardial infarction (TIMI) 3 flow in the infarct-related artery without any complication.

The study protocol received approval by the local Ethics Committees.

### Outcomes

The primary outcome was a composite of 30-day all-cause mortality or complications requiring ICU admission occurring during the index hospitalization. Pre-specified ICU-level complications included: acute heart failure with pulmonary oedema, cardiogenic shock requiring pharmacological or mechanical support, malignant arrhythmias (e.g., ventricular fibrillation, sustained ventricular tachycardia, high-grade atrioventricular block, or asystole), need for invasive mechanical ventilation, acute and subacute stent thrombosis, and reinfarction. The secondary endpoint was all-cause mortality at 2 years.

### Statistical Analysis

Categorical variables are reported as counts and percentages, and compared using chi-square test. Continuous variables are presented as mean ± standard deviation (SD) or median [interquartile range, IQR], and compared using Student’s t-test or Mann-Whitney U test, as appropriate. To ensure robust temporal validation, the cohort was split into a derivation (patients undergoing pPCI on Tuesday, Wednesday, Friday, Saturday and Sunday n=3,649) and a validation set (pPCI on Monday, Thursday; n=1,444). Variables associated with the primary outcome in univariate logistic regression (p<0.05) were entered into a multivariable model. To maintain immediate clinical applicability of the score, variables not available at the time of PCI completion (e.g., left ventricular ejection fraction, haemoglobin, creatinine) were excluded from the model. Independent predictors were weighted based on adjusted odds ratios, rounded to the nearest integer, and summed into the Codi-IAM Score. Model discrimination was assessed using the area under the receiver-operating characteristic curve (AUC), and calibration was evaluated by comparing observed and predicted event rates across risk quintiles. Risk categories were predefined as follows: very low (0 points), low (1–4), moderate (5–9), high (10–14), and very high (>14). Kaplan–Meier survival curves were generated and compared using log-rank tests at 30-day and 2-year follow-up. Additional exploratory analyses included a multivariable model for predictors of acute stent thrombosis and a landmark analysis evaluating primary events occurring beyond the first 6 hours post-pPCI. All statistical analyses were performed using SPSS version 25 (IBM Corp., Armonk, NY). A two-sided p-value <0.05 was considered statistically significant.

## RESULTS

### Baseline characteristics

A total of 5,093 STEMI patients undergoing primary PCI were included in the analysis. The mean age was 63.0 ± 12.9 years. Women represented 21% of the cohort (n=1,101). Baseline characteristics were well balanced between the derivation (n=3,649) and validation (n=1,444) cohorts (**Table 1**). Overall, 49.3% of patients had hypertension, 23% had diabetes mellitus, and 44% were current smokers. Dyslipidemia and chronic renal failure were slightly more prevalent in the derivation cohort compared to the validation cohort (42% vs 39%, p=0.037; and 5.0% vs 3.5%, p=0.025). Approximately 94% of patients were triaged via EMS or referring hospitals in both groups, with no differences. The median symptom onset-to-balloon time was ∼3 hours, and the median interval between the first medical contact-to-ECG was 8 minutes [IQR 5–17]. (**Supplementary Table 1**) Radial artery access was used in 93.7% of the procedures, and stents were implanted in the 93.8% of cases (**Supplementary Table 2**). At the time of presentation, 998 patients (19.6%) were in Killip class > I; 175 (3.4%) had angiographical evidence of left main disease and 672 (13%) had three-vessel disease. Arrhythmic events prior to PCI completion occurred in 1,213 patients (23.8%), predominantly accelerated idioventricular rhythm (4.0%), sustained ventricular fibrillation (3.0%), and ventricular tachycardia (2.0%) (**Supplementary Table 2**).

**Table 1.**
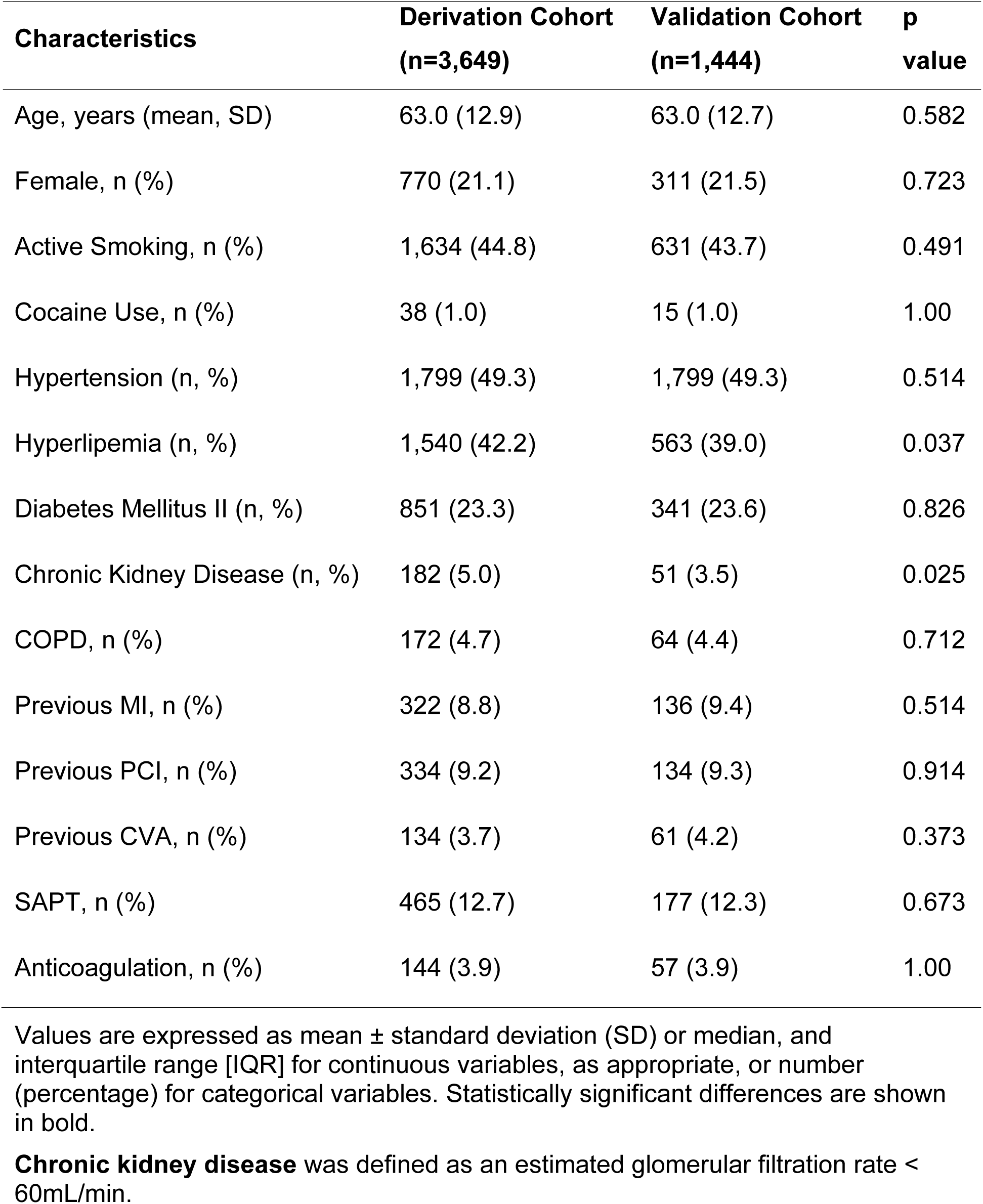
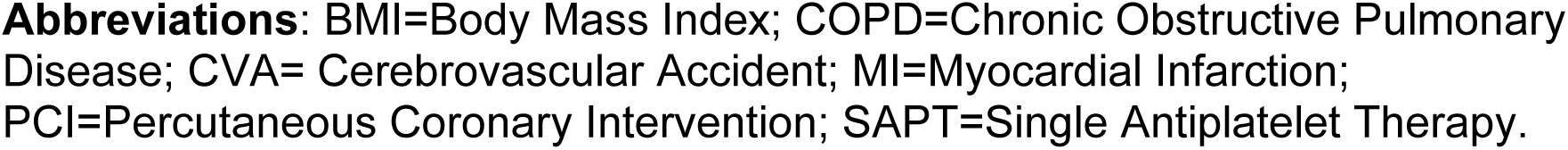
Baseline clinical characteristics of the overall study population and comparison between derivation and validation cohorts.

| Characteristics | Derivation Cohort<br>(n=3,649) | Validation Cohort<br>(n=1,444) | p<br>value |
| --- | --- | --- | --- |
| Age, years (mean, SD) | 63.0 (12.9) | 63.0 (12.7) | 0.582 |
| Female, n (%) | 770 (21.1) | 311 (21.5) | 0.723 |
| Active Smoking, n (%) | 1,634 (44.8) | 631 (43.7) | 0.491 |
| Cocaine Use, n (%) | 38 (1.0) | 15 (1.0) | 1.00 |
| Hypertension (n, %) | 1,799 (49.3) | 1,799 (49.3) | 0.514 |
| Hyperlipemia (n, %) | 1,540 (42.2) | 563 (39.0) | 0.037 |
| Diabetes Mellitus II (n, %) | 851 (23.3) | 341 (23.6) | 0.826 |
| Chronic Kidney Disease (n, %) | 182 (5.0) | 51 (3.5) | 0.025 |
| COPD, n (%) | 172 (4.7) | 64 (4.4) | 0.712 |
| Previous MI, n (%) | 322 (8.8) | 136 (9.4) | 0.514 |
| Previous PCI, n (%) | 334 (9.2) | 134 (9.3) | 0.914 |
| Previous CVA, n (%) | 134 (3.7) | 61 (4.2) | 0.373 |
| SAPT, n (%) | 465 (12.7) | 177 (12.3) | 0.673 |
| Anticoagulation, n (%) | 144 (3.9) | 57 (3.9) | 1.00 |

Values are expressed as mean ± standard deviation (SD) or median, and interquartile range [IQR] for continuous variables, as appropriate, or number (percentage) for categorical variables. Statistically significant differences are shown in bold.
**Chronic kidney disease** was defined as an estimated glomerular filtration rate < 60mL/min.
**Abbreviations:** BMI=Body Mass Index; COPD=Chronic Obstructive Pulmonary Disease; CVA= Cerebrovascular Accident; MI=Myocardial Infarction; PCI=Percutaneous Coronary Intervention; SAPT=Single Antiplatelet Therapy.

### Primary and secondary outcomes

The primary composite endpoint occurred in 1,501 patients (29.5%). 30-day all-cause mortality occurred in 5.5% of the patients, with no significant difference between the derivation (5.7%) and validation (5.1%) cohorts (p=0.46). (**Central Illustration**) In-hospital complications requiring ICU care since the first medical contact occurred in 17.8% of patients (18.1% derivation vs 16.9% validation, p=0.31). Major complications included cardiogenic shock (4.8%), malignant ventricular arrhythmias (3.1%), and the need for invasive mechanical ventilation (2.8%). Acute stent thrombosis occurred in 38 patients (0.7%), with 95% of cases occurring within 3 hours post-PCI. Additional in-hospital outcomes are summarized in **Table 2**.

**Table 2.** Complications occurred during hospitalization (in-hospital), and 30-day mortality rates were assessed in the overall cohort, as well as in the derivation and validation cohorts.

| **Characteristics** | **Overall Cohort**<br>**(n=5,093)** | **Derivation Cohort**<br>**(n=3,649)** | **Validation Cohort**<br>**(n=1,444)** | **p value** |
| --- | --- | --- | --- | --- |
| 30-day death | 282 (5.5) | 208 (5.7) | 74 (5.1) | 0.455 |
| Acute Pulmonary Edema | 88 (1.7) | 70 (1.9) | 18 (1.2) | 0.012 |
| Cardiogenic Shock | 242 (4.8) | 182 (5.0) | 60 (4.2) | 0.215 |
| Mechanical Ventilation, n (%) | 142 (2.8) | 101 (2.8) | 41 (2.8) | 0.925 |
| Malignant Arrhythmias, n (%) |  |  |  |  |
| - Asystole | 159 (3.1) | 113 (3.1) | 46 (3.2) | 0.858 |
| - Ventricular Fibrillation | 129 (2.5) | 97 (2.7) | 32 (2.2) | 0.429 |
| - Ventricular Tachycardia | 40 (0.8) | 24 (0.7) | 16 (1.1) | 0.113 |
| - Atrio-Ventricular Block | 137 (2.7) | 102 (2.8) | 35 (2.4) | 0.502 |
| - Atrial Fibrillation | 127 (2.5) | 92 (2.5) | 35 (2.4) | 0.921 |
| - Others | 64 (1.3) | 45 (1.2) | 19 (1.3) | 0.782 |
| Acute Stent-Thrombosis, n (%) | 38 (0.7) | 26 (0.7) | 12 (0.8) | 0.718 |
| Re-infarction (<24h) n (%) | 17 (0.3) | 12 (0.3) | 5 (0.3) | 1.0 |
| Mechanical Complications, n (%) | 36 (0.7) | 27 (0.7) | 9 (0.6) | 0.715 |
| Overall, in-hospital Complications, n (%) | 906 (17.8) | 662 (18.1) | 244 (16.9) | 0.310 |

Values are expressed as number (percentage).

### Development and validation of the Codi-IAM Score

Seven independent predictors of the primary endpoint were identified in multivariable logistic regression (all p<0.05): age >75 years (OR 1.79), Killip class > I (OR 6.36), malignant arrhythmia before PCI completion (OR 9.49), left main disease (OR 2.04), three-vessel disease (OR 1.36), non-radial access (OR 1.79), and unsuccessful PCI (final TIMI flow < 3; OR 1.90). Each predictor was weighted by its odds ratio to create the Codi-IAM Score (range: 0–24 points) (**Supplementary Table 3**). Risk categories were defined as very low (0 points), low (1–4), moderate (5–9), high (10–14), and very high (>14). The model demonstrated excellent discrimination (c-statistic: 0.87 [95% CI: 0.85–0.89] in the derivation cohort, and 0.86 [95% CI: 0.83–0.89] in the validation cohort; both p<0.001), and strong calibration. (**Central Illustration**)

### Outcomes stratified by risk category

Among patients in the very low-risk (score 0; 44.3% of total), there were no deaths within 72 hours post-pPCI, and 30-day mortality was 0.2% (n=5). Only one death that occurred seven days post-procedure was attributed to the index myocardial infarction and interpreted as cardiac death. The remaining four deaths were non-cardiac (major bleeding due to massive epistaxis, pneumonia, stroke, metastatic cancer). (**Figure 1**) In-hospital complications occurred in 1.7% of very low-risk patients, including cardiogenic shock (1 patient, 0.1%) and malignant arrhythmias (1 patient, 0.1%), both in the context of acute stent thrombosis. Most episodes of ventricular tachycardia were non-sustained and did not require cardioversion (**Table 3**). Acute stent thrombosis occurred in 13 patients (0.6%) in the very low-risk group. Landmark analysis showed that 12 of these events occurred within the first hour after pPCI, while only one (0.02%) occurred beyond 6 hours, specifically at 72 hours, highlighting the immediate post-PCI period as the highest-risk window for this complication. Additional multivariate analysis of predictors of acute stent thrombosis is provided in the **Supplementary Materials** (**Supplementary Table 4**).

**Figure 1.**
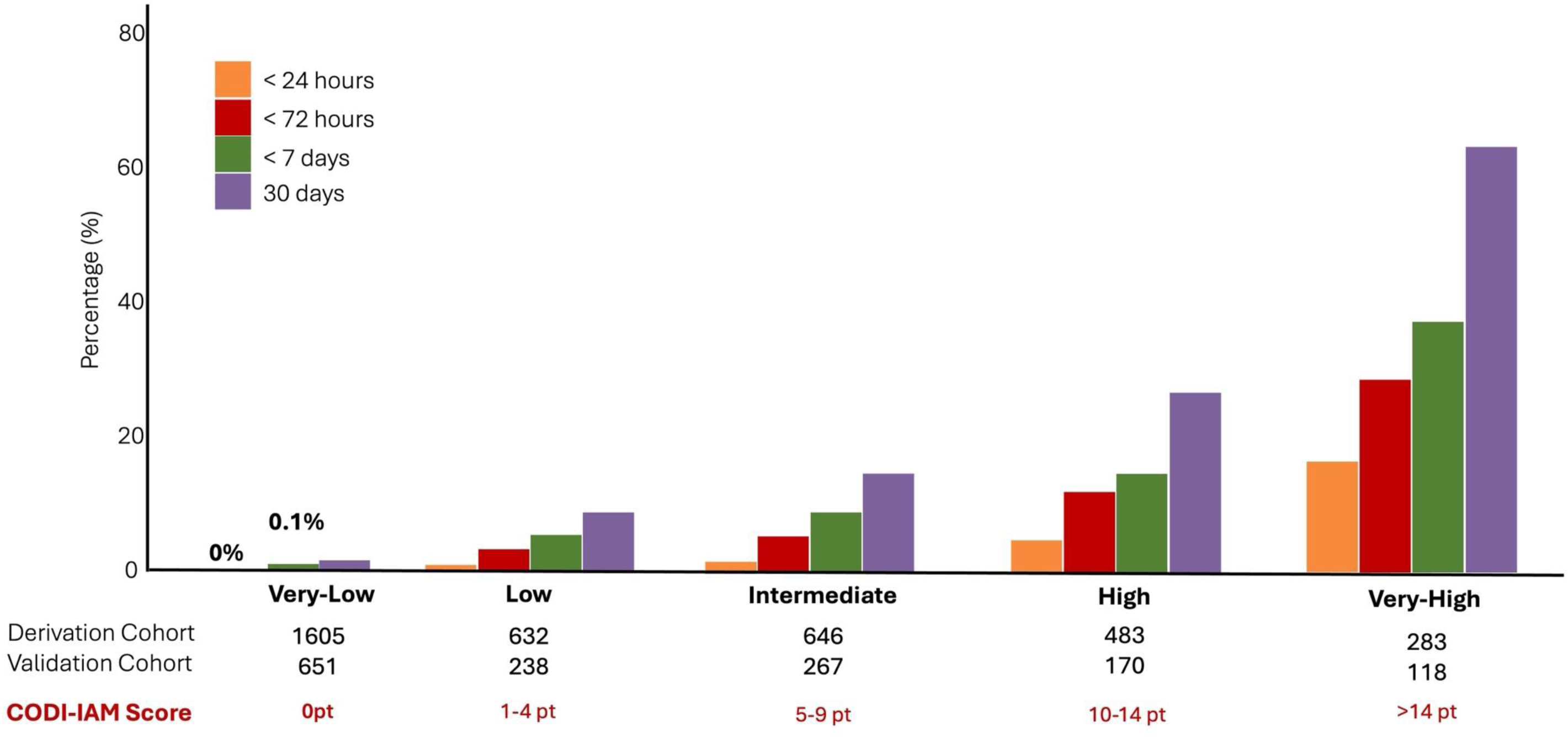
Early and 30-Day Mortality by Risk Category According to Codi-IAM Score. The diagram shows mortality at 24 hours, 3 days, 7 days, and 30 days stratified by risk categories defined by the Codi IAM-Code Score: very low-risk (score 0), low-risk (1–4 points), intermediate-risk (5–9 points), high-risk (10–14 points), and very high-risk (>14 points). Patients classified as very low-risk (score 0) exhibited no deaths within the first 72 hours and a 30-day mortality of only 0.2%. Mortality increased progressively across higher risk categories, highlighting the robust stratification capacity of the score. This figure supports the use of the Codi-IAM Score for real-time post-pPCI risk assessment and clinical decision-making regarding ICU discharge.

**Table 3.**
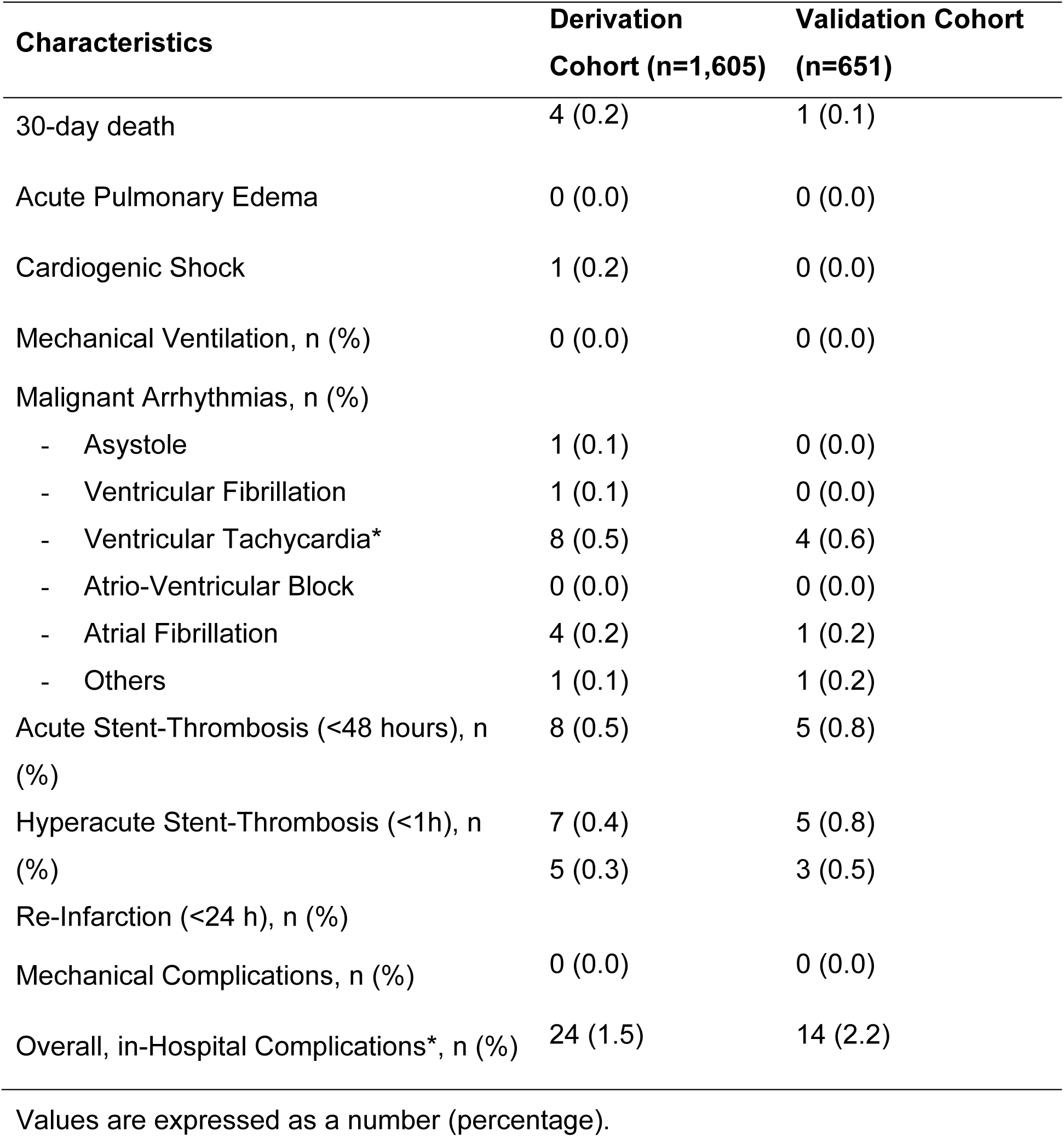
Clinical outcomes among patients classified as very low-risk (Codi-IAM Score = 0).

Patients with any score ≥1 had a significantly higher 30-day mortality rate compared to those with a score of 0 (7.3% vs 0.2%, p<0.001). Kaplan–Meier analysis shows 2-year survival exceeding 98.4% in the very low-risk group, clearly separated from all higher-risk strata (p<0.001) (**Figure 2**).

**Figure 2.**
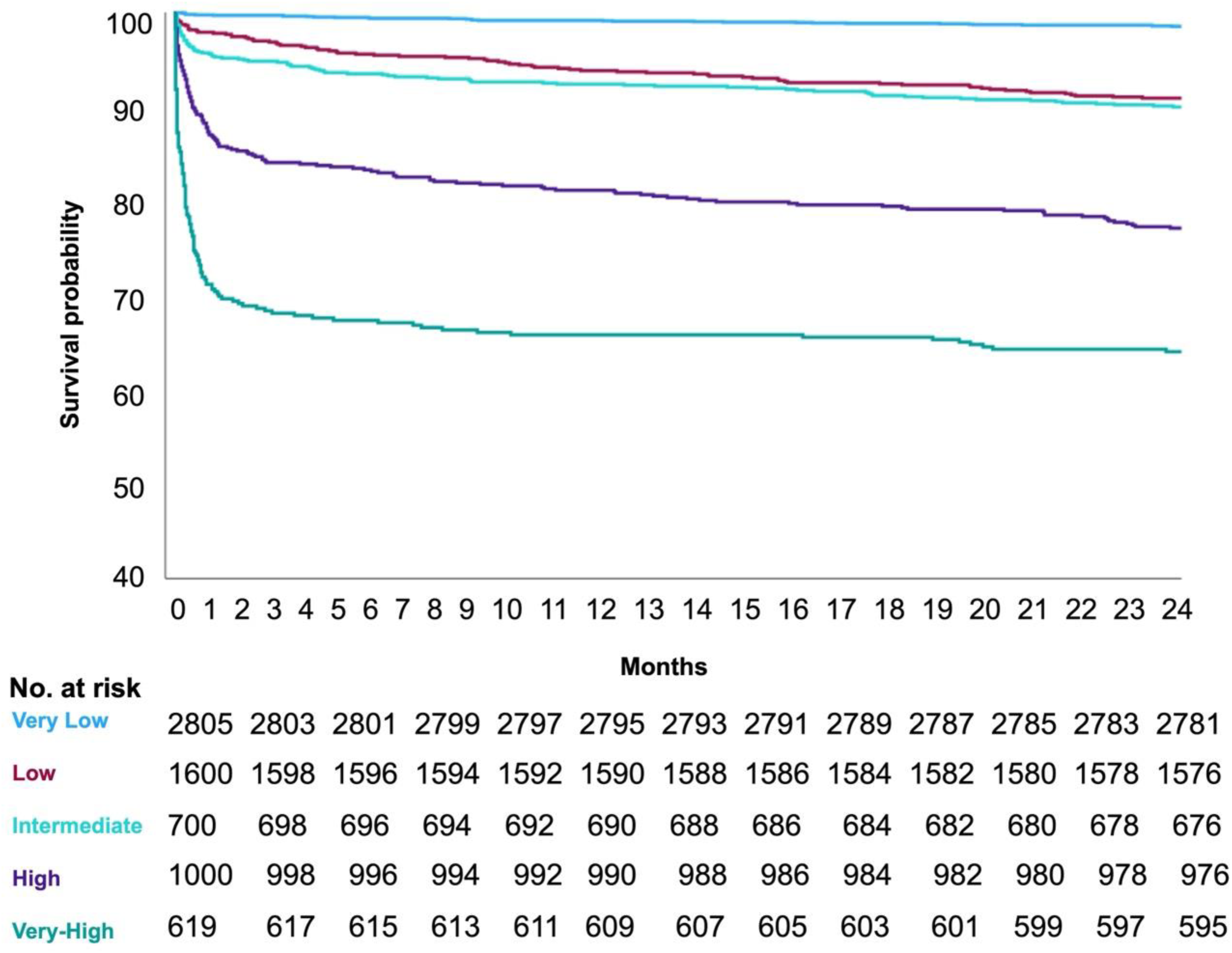
Survival curves at two-year follow-up. Kaplan-Meier curves show two-year mortality stratified by Codi-IAM Score. Patients classified as very low risk (Codi-IAM score = 0) exhibited consistently low mortality throughout the follow-up period.

## DISCUSSION

This prospective, multicenter study provides robust evidence supporting the efficacy of the Codi-IAM Score in identifying STEMI patients at very low risk of early adverse events, enabling safe reductions in ICU stays without compromising patient outcomes. The key findings can be summarized as follows (i) a simple set of clinical and procedural characteristics, including age <75 years, Killip class I, absence of peri-procedural malignant arrhythmias, absence of left main or three-vessel coronary artery disease, radial access, and successful PCI, was associated with a negligible risk of 30-day mortality or ICU-requiring complications; ii) the score demonstrated excellent discriminatory capacity (AUC=0.87 in the derivation cohort, AUC=0.86 in the validation cohort) with nearly half of the patients classified as very low risk, experiencing minimal adverse events. Our findings significantly expand our previous single-center data, confirming that appropriately selected STEMI patients may safely undergo early discharge.(9) In addition, the results are consistent with those from Rathod et al., who demonstrated that low-risk STEMI patients discharged within 48 hours had comparable or superior outcomes compared to those with conventional length of stay (1, 8). The Codi-IAM Score provides a pragmatic alternative to established risk models (e.g., Zwolle, PAMI-II, CADILLAC scores), by exclusively incorporating variables immediately available post-PCI, reflecting modern interventional practices including radial access, contemporary stenting techniques, and guideline-based secondary prevention strategies.(7, 11, 12, 13) While core clinical predictors (e.g., advanced age, higher Killip class) are consistent with prior literature (14) and emphasize their clinical significance as determinants of short-term prognosis, the contemporary real-life derivation, the only inclusion of clinical, and angiographic data resulting in a point-of-care applicability enhance the value of our model. Killip class > I emerged as a strong predictor in the model. This is in line with historical and contemporary data showing that clinical signs of heart failure at presentation are tightly linked to short-term mortality, reflecting both left ventricular dysfunction and elevated filling pressures.(15) The hemodynamic instability captured by Killip class ≥II remains a cornerstone of early risk stratification in STEMI, even in the era of timely reperfusion.(14, 16) Age over 75 years in our score was associated with a significantly increased risk of adverse outcomes, a finding consistent with previous literature highlighting the cumulative impact of comorbidities, frailty, and impaired cardiovascular reserve in elderly patients.(17, 18) Advanced age also correlates with diminished tolerance to ischemia and a higher likelihood of complications such as bleeding or contrast-induced nephropathy, all contributing to early mortality after STEMI. Other than age and Killip class, peri-procedural malignant arrhythmias significantly influence early outcomes by identifying patients with transient but critical electrical instability during acute ischemia, as previously demonstrated.(19, 20) From an anatomical perspective, complex coronary anatomy (e.g., left main, multivessel disease) further delineates a patient subgroup characterized by greater myocardial territory at risk, hemodynamic instability, and heightened risk of post-procedural complications.(21, 22) In addition, final TIMI flow <3 also remains a potent angiographic predictor, emphasizing the critical importance of optimal epicardial and microvascular reperfusion for reducing infarct size, arrhythmic risk, and mechanical complications.(23, 24) The alignment with prior evidence gives consistency and external validity to the model, while their combined performance supports a safe and reproducible early discharge strategy.

Importantly, the robust methodology, the comprehensive real-world design, and the rigorous quality control within the Catalonia Codi-IAM network enhance the validity and applicability of our findings, demonstrating the key value of standardized regional care systems.

Taken together, the guidance of an early ICU discharge, through the application of the Codi-IAM Score may have significant positive implications for both patients and hospital. It may increase healthcare efficiency, reduced costs, improved resource allocation, and increased patients’ compliance and satisfaction. Of note, the excellent short-term safety profile of the very low-risk group was extended 2 years, suggesting the potential utility of this score also for longitudinal planning and follow-up intensity calibration.

## STUDY LIMITATIONS

Limitations of our study include the modest predictive value of the Codi-IAM score for acute stent thrombosis. Although rare (0.7%), >95% of the events occurred within the first hour post-PCI, including all but one in the very low-risk group (0.02%). This tight temporal profile suggests that even if the score does not fully capture the risk of this event, the occurrence within a short time-window allows for a targeted monitoring strategy, supporting the concept that clinical observation during the first 6 hours remains sufficient for safely detecting this complication, even in low-risk patients. Furthermore, our study includes its derivation and validation within a structured regional healthcare network, cautioning against direct extrapolation to less structured or resource-limited environments. Finally, although our data collection was comprehensive and quality-controlled, residual confounding is possible and inherent in any observational study.

## CONCLUSIONS

In a contemporary, high-volume STEMI network, our study strongly supports the clinical utility of the Codi-IAM Score, facilitating accurate, real-time identification of STEMI patients at very low risk following pPCI. Implementing this score in clinical practice can safely guide ICU discharge decisions, optimize resource utilization, and maintain clinical safety, reinforcing the feasibility and benefits of structured early discharge pathways.

## Data Availability

The data that support the findings of this study are available from the corresponding author upon reasonable request. The data are not publicly available due to restrictions related to patient privacy and institutional regulations.

## Acknowledgements

None.

## Sources of Funding

None.

## Disclosures

None of the Authors has a conflict of interest to declare.

## Nonstandard Abbreviations and Acronyms

AMI: Acute Myocardial Infarction
ECG: Electrocardiogram
EMS: Emergency Medical Services
FMC: First Medical Contact
ICU: Intensive Care Unit
PCI: Percutaneous Coronary Intervention
pPCI: Primary Percutaneous Coronary Intervention
STEMI: ST-Elevation Myocardial Infarction
TIMI: Thrombolysis in Myocardial Infarction

---

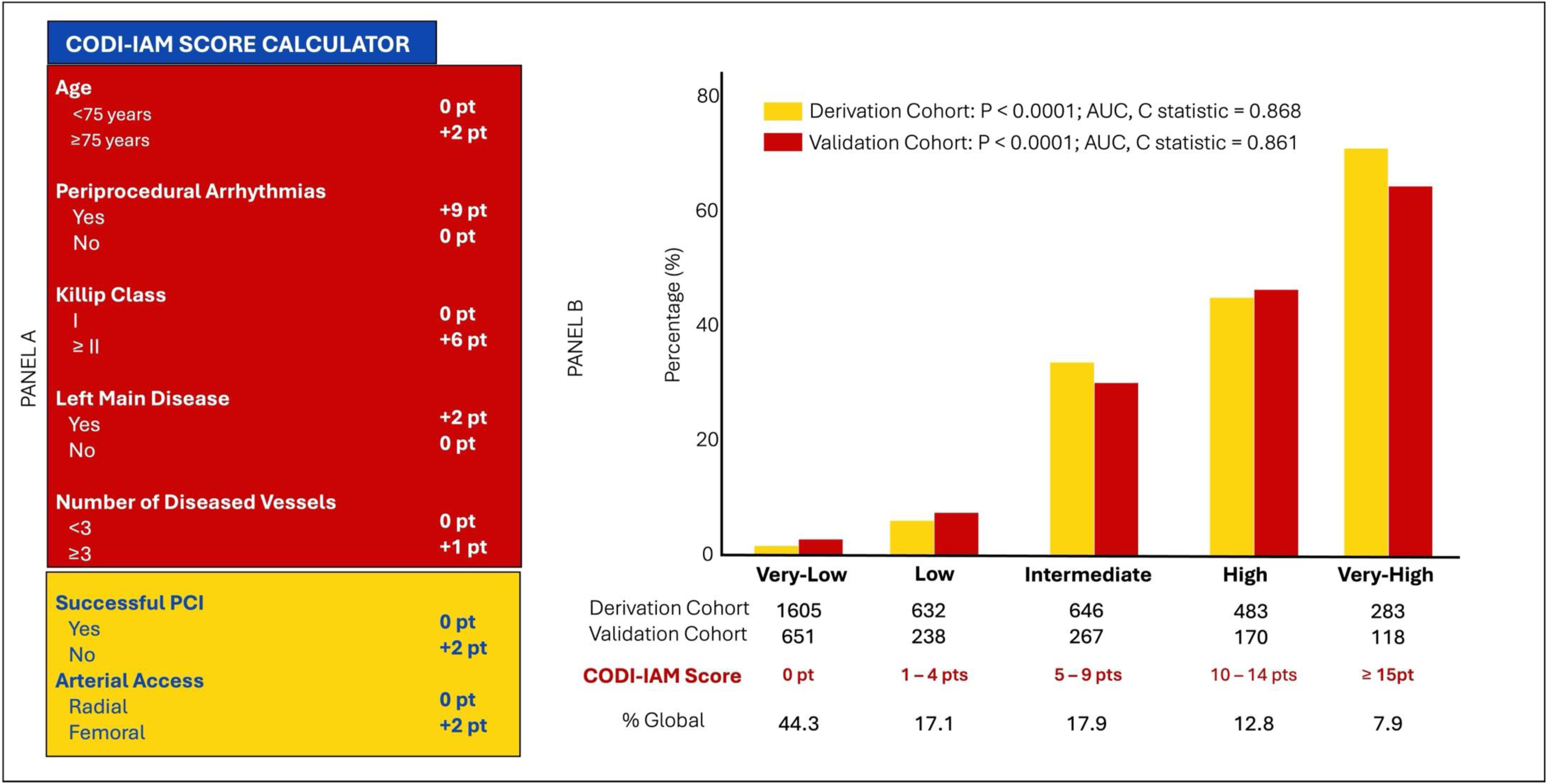
Central Illustration. **Development and performance of the Codi-IAM Score.** The **Panel A** shows the clinical and procedural variables independently associated with the composite endpoint of 30-day all-cause mortality or ICU-requiring complications that were weighted according to their odds ratios to construct the Codi-IAM Score (range 0–24 points). The **Panel B** shows the distribution of 30-day event rates across predefined risk categories (very-low, low, moderate, high, and very high) in both the derivation and validation cohorts. The score demonstrated excellent discrimination and calibration (Derivation Cohort: P < 0.0001; AUC, C statistic = 0.868; Validation Cohort: P < 0.0001; AUC, C statistic = 0.861), reliably stratifying STEMI patients across their risk spectrum.

## Notes

### Competing Interest Statement

The authors have declared no competing interest.

### Author Declarations

The study was approved by the Institutional Review Board (IRB) / Ethics Committee of the Hospital Clinic of Barcelona, in accordance with the Declaration of Helsinki.

## REFERENCES

1. Rathod KS, Comer K, Casey-Gillman O, Moore L, Mills G, Ferguson G, et al. Early Hospital Discharge Following PCI for Patients With STEMI. J Am Coll Cardiol. 2021;78(25):2550–60.

2. Yndigegn T, Gilje P, Dankiewicz J, Mokhtari A, Isma N, Holmqvist J, et al. Safety of early hospital discharge following admission with ST-elevation myocardial infarction treated with percutaneous coronary intervention: a nationwide cohort study. EuroIntervention. 2022;17(13):1091–9.

3. Gong W, Li A, Ai H, Shi H, Wang X, Nie S. Safety of early discharge after primary angioplasty in low-risk patients with ST-segment elevation myocardial infarction: A meta-analysis of randomised controlled trials. Eur J Prev Cardiol. 2018;25(8):807–15.

4. Byrne RA, Rossello X, Coughlan JJ, Barbato E, Berry C, Chieffo A, et al. 2023 ESC Guidelines for the management of acute coronary syndromes. Eur Heart J. 2023;44(38):3720–826.

5. Rao SV, O’Donoghue ML, Ruel M, Rab T, Tamis-Holland JE, Alexander JH, et al. 2025 ACC/AHA/ACEP/NAEMSP/SCAI Guideline for the Management of Patients With Acute Coronary Syndromes: A Report of the American College of Cardiology/American Heart Association Joint Committee on Clinical Practice Guidelines. J Am Coll Cardiol. 2025.

6. De Luca G, Suryapranata H, van ‘t Hof AW, de Boer MJ, Hoorntje JC, Dambrink JH, et al. Prognostic assessment of patients with acute myocardial infarction treated with primary angioplasty: implications for early discharge. Circulation. 2004;109(22):2737–43.

7. Grines CL, Marsalese DL, Brodie B, Griffin J, Donohue B, Costantini CR, et al. Safety and cost-effectiveness of early discharge after primary angioplasty in low risk patients with acute myocardial infarction. PAMI-II Investigators. Primary Angioplasty in Myocardial Infarction. J Am Coll Cardiol. 1998;31(5):967–72.

8. Rathod KS, Comer K, Casey-Gillman O, Moore L, Antoniou S, Fhadil S, et al. Cost-Effectiveness of Early Discharge (<48 Hours) for Low-Risk Patients Following PPCI for STEMI. JACC Cardiovasc Interv. 2025;18(12):1499–509.

9. Altisent OA, Carrillo X, Puri R, Bayes-Genis A. Identifying very low-risk STEMI patients for early ICU discharge in the COVID-19 era. Clin Res Cardiol. 2020;109(12):1582–4.

10. Regueiro A, Tresserras R, Goicolea J, Fernandez-Ortiz A, Macaya C, Sabate M. Primary percutaneous coronary intervention: models of intervention in Spain. EuroIntervention. 2012;8 Suppl P:P90–3.

11. Wilson RS, Malamas P, Dembo B, Lall SK, Zaman N, Peterson BR. The CADILLAC risk score accurately identifies patients at low risk for in-hospital mortality and adverse cardiovascular events following ST elevation myocardial infarction. BMC Cardiovasc Disord. 2021;21(1):533.

12. Valgimigli M, Gagnor A, Calabro P, Frigoli E, Leonardi S, Zaro T, et al. Radial versus femoral access in patients with acute coronary syndromes undergoing invasive management: a randomised multicentre trial. Lancet. 2015;385(9986):2465–76.

13. Gargiulo G, Giacoppo D, Jolly SS, Cairns J, Le May M, Bernat I, et al. Effects on Mortality and Major Bleeding of Radial Versus Femoral Artery Access for Coronary Angiography or Percutaneous Coronary Intervention: Meta-Analysis of Individual Patient Data From 7 Multicenter Randomized Clinical Trials. Circulation. 2022;146(18):1329–43.

14. Del Buono MG, Montone RA, Rinaldi R, Gurgoglione FL, Meucci MC, Camilli M, et al. Clinical predictors and prognostic role of high Killip class in patients with a first episode of anterior ST-segment elevation acute myocardial infarction. J Cardiovasc Med (Hagerstown). 2021;22(7):530–8.

15. Vicent L, Velasquez-Rodriguez J, Valero-Masa MJ, Diez-Delhoyo F, Gonzalez-Saldivar H, Bruna V, et al. Predictors of high Killip class after ST segment elevation myocardial infarction in the era of primary reperfusion. Int J Cardiol. 2017;248:46–50.

16. de Carvalho LP, Gao F, Chen Q, Sim LL, Koh TH, Foo D, et al. Long-term prognosis and risk heterogeneity of heart failure complicating acute myocardial infarction. Am J Cardiol. 2015;115(7):872–8.

17. Castello R, Alegria E, Merino A, Malpartida F, Martinez-Caro D. Effect of age on long-term prognosis of patients with myocardial infarction. Int J Cardiol. 1988;20(2):221–30.

18. Nishihira K, Kuriyama N, Kadooka K, Honda Y, Yamamoto K, Nishino S, et al. Outcomes of Elderly Patients With Acute Myocardial Infarction and Heart Failure Who Undergo Percutaneous Coronary Intervention. Circ Rep. 2022;4(10):474–81.

19. Mehta RH, Starr AZ, Lopes RD, Hochman JS, Widimsky P, Pieper KS, et al. Incidence of and outcomes associated with ventricular tachycardia or fibrillation in patients undergoing primary percutaneous coronary intervention. JAMA. 2009;301(17):1779–89.

20. Demidova MM, Holmqvist F, Erlinge D, Platonov PG. Ventricular arrhythmias during ST-segment elevation myocardial infarction and arrhythmic complications during recurrent ischaemic events. Eur Heart J. 2024;45(5):393–5.

21. Faro DC, Laudani C, Agnello FG, Ammirabile N, Finocchiaro S, Legnazzi M, et al. Complete Percutaneous Coronary Revascularization in Acute Coronary Syndromes With Multivessel Coronary Disease: A Systematic Review. JACC Cardiovasc Interv. 2023;16(19):2347–64.

22. Laudani C, Occhipinti G, Greco A, Spagnolo M, Giacoppo D, Capodanno D. Completeness, timing, and guidance of percutaneous coronary intervention for myocardial infarction and multivessel disease: a systematic review and network meta-analysis. EuroIntervention. 2025;21(4):e203–e16.

23. Vichova T, Maly M, Ulman J, Motovska Z. Mortality in patients with TIMI 3 flow after PCI in relation to time delay to reperfusion. Biomed Pap Med Fac Univ Palacky Olomouc Czech Repub. 2016;160(1):118–24.

24. Stone GW, Cox D, Garcia E, Brodie BR, Morice MC, Griffin J, et al. Normal flow (TIMI-3) before mechanical reperfusion therapy is an independent determinant of survival in acute myocardial infarction: analysis from the primary angioplasty in myocardial infarction trials. Circulation. 2001;104(6):636–41.

